# ClimateMind50+: Development and validation through cognitive interviews of a questionnaire to measure climate change knowledge, concerns, and actions in older adults

**DOI:** 10.1101/2024.12.02.24318302

**Authors:** Paola Zaninotto, Yu-Tzu Wu, Matthew Prina

**Author notes:** **Corresponding author** Professor Paola Zaninotto Department of Epidemiology and Public Health, University College London 1-19 Torrington Place, London WC1E 7HB, United Kingdom.

## Abstract

**Background and aim:** Older adults are particularly vulnerable to climate-related hazards such as extreme heat, flooding, and severe storms, yet their perspectives and contributions to climate resilience remain underrepresented in research. The ClimateMind50+ questionnaire is a tailored instrument to assess the knowledge, concerns, preparedness, behaviours, and involvement of individuals aged 50 and above.

**Methods:** The systematic development of the ClimateMind50+ involved rigorous cognitive testing with 15 diverse participants, ensuring clarity, accessibility, and relevance. Review by experts helped to refine its content and ensure its comprehensiveness. Designed for versatility in administration (face-to-face, via telephone, or self-completion) to facilitate its integration across various research contexts.

**Results:** Experts’ input led to refinements to ensure the questionnaire effectively captures older adults knowledge, concerns, preparedness, and involvement in climate action while making it more accessible. Cognitive testing highlighted the need for clear wording, simplified response scales, and age-appropriate framing of questions. For instance, questions on climate preparedness and sustainable practices were refined to capture lifetime actions (“ever”) rather than limited timeframes, enhancing their relevance for older respondents.

**Conclusions:** By providing nuanced insights into the experiences of older adults and their potential contributions to climate mitigation and adaptation, the ClimateMind50+ offers a robust foundation for climate change research among older people. Its deployment can support policymaking and community initiatives aimed at reducing climate risks while promoting sustainable and healthy aging practices. This innovative tool underscores the importance of amplifying the voices of older adults in climate discourse and harnessing their capacities for fostering resilience.

## Introduction

The escalating challenges posed by climate change demand a nuanced understanding of its impacts on vulnerable populations, particularly older adults.^1^ Older individuals are disproportionately affected by climate-related phenomena such as extreme heat, flooding, and severe weather, which exacerbate existing health conditions and introduce unique vulnerabilities.^1^ Yet, despite their heightened risk, this demographic is often underrepresented in studies addressing the relationship of climate change and healthy aging. Understanding how older adults perceive and respond to climate risks is crucial for developing targeted interventions that enhance their resilience and well-being.

We previously conducted a systematic mapping of aging studies^2^ which revealed significant gaps in the integration of climate-related measures. While recently a measure has been developed to examine the perceived negative and positive contributions of older people to climate change,^3^ few studies employ questionnaires tailored to assess older adults’ concerns,^4^ perceptions, preparedness, and behaviours^3,4^ regarding climate change. Existing tools are either outdated or not specifically designed for aging populations, limiting their capacity to capture the unique experiences and potential contributions of older people.^4^

To address these gaps, this article introduces and describes in detail the development of a new, concise questionnaire specifically designed for individuals aged 50 and above, created for administration within the English Longitudinal Study of Ageing (ELSA)^5^ and intended to be made available for use in other ageing studies. The primary objective of this research is to develop a tool measuring key aspects of climate-related knowledge, concerns, and adaptive behaviours among older adults and to establish its content validity among older adults. This tool aims to address critical dimensions such as climate worry, preparedness for extreme events, and engagement in climate action. By focussing on the experiences and perspectives of older adults, this questionnaire not only fills an empirical gap but also challenges prevailing stereotypes that marginalize this group as passive or disengaged in climate discourse. Moreover, it recognizes the untapped potential of older individuals as active participants in fostering sustainable practices and mitigating climate change. ^6,7^

The questionnaire will generate valuable age-specific data on climate change attitudes, behaviours, worries, and engagement among older adults, which in turn will support the development of targeted policies and strategies to protect and empower older populations in the face of a changing climate.

## Methods

### Identification of key aspects of climate-related issues in older people

We reviewed the 153 ageing studies in three cohort consortiums (Integrative Analysis of Longitudinal Studies of Ageing and Dementia [IALSA],^8^ Ageing Trajectories of Health: Longitudinal Opportunities and Synergies [ATHLOS],^9^ and Gateway to Global Ageing Data^10^) and identified survey questions related to climate change.^2^ Furthermore, we carried out additional searches to identify surveys and questionnaires used in general populations (of all ages) and government reports, with a particular focus on data from the UK and Europe.

Based on the identified survey questions, the topics were categorised into four groups:

1. Attitudes: Understanding Society - New module on environmental attitudes (2022),^11^ Solastalgia,^12^ Office for National Statistics Public attitudes to the environment and the impact of climate change, Great Britain (2021),^13^ Department for Energy Security and Net Zero (DESNZ) and Department for Business, Energy and Industrial Strategy (BEIS) Public Attitude Tracker,^14^ and Climate Change in the Irish Mind (CCIM);^15^
2. Behaviours: General Ecological Behaviour Scale,^4^ Sustainable consumption behaviour,^16^ Understanding Society - Environmental behaviours,^11^ and CCIM;^15^
3. Worries: Climate change anxiety scale,^17^ DESNZ/BEIS Public Attitude,^14^ and CCIM;^15^
4. Beliefs, perceptions and knowledge: Centre for Climate Change and Social Transformations (CAST),^18,19^ Climate change and Net Zero: Public Awareness and Perceptions (2021),^20^ European Perceptions of Climate Change (2016),^21^ British Public Perceptions of Climate Risk, Adaptation Options and Resilience (RESiL RISK) (2019),^22^ CCIM,^15^ and DESNZ/BEIS Public Attitude Tracker.^14^

### Expert panel review

The questions were meticulously reviewed and refined, with new ones developed to ensure their suitability for administration to older adults. This process involved consultation with experts in survey methods, climate change, gerontology, aging, social policy, and epidemiology. We carefully examined individual questions from existing surveys, selecting those most relevant to older individuals. The selected questions underwent thorough discussion, with modifications made to align with the requirements for administration in ageing studies. The questionnaire was specifically designed to address key challenges older adults face in relation to climate change, including their increased vulnerability to extreme weather events, preparedness for climate-related risks, and engagement in mitigation and adaptation efforts. Multiple iterations were conducted to achieve a consensus on a finalized set of questions (Figure 1).

**Figure 1.**
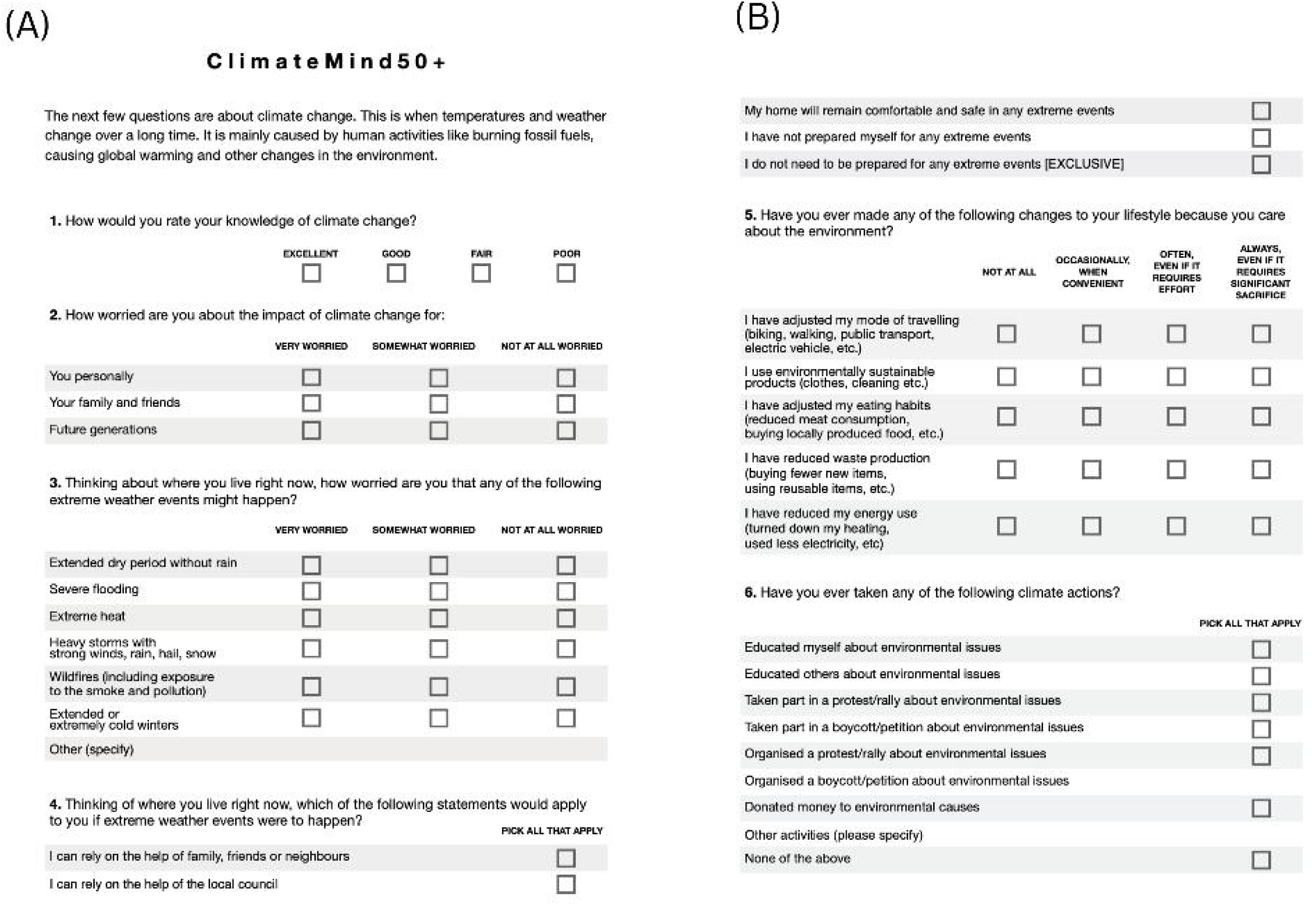
ClimateMind50+ questionnaire

### Cognitive testing interviews

We then conducted cognitive testing interviews to explore how participants mentally processed the ClimateMind50+ survey questions, aiming to improve the clarity and effectiveness of survey designs. These interviews played a crucial role in understanding whether participants could interpret questions accurately, recall relevant information, and provide thoughtful responses. Rather than focusing on the specific answers given, the process investigated how individuals engaged with the survey itself, identifying potential barriers to comprehension or response accuracy.

Each interview was conducted one-on-one to allow for an in-depth exploration of the participant’s thought process. Participants were encouraged to articulate their understanding of the questions, describe how they recalled the necessary information, and explain their decision-making strategies when selecting answers. Feedback was recorded during each individual interview. Additionally, the interviews assessed their comfort level in responding to certain questions and their confidence in the accuracy of their answers. This approach provided valuable insights into how survey questions might be misunderstood or misinterpreted, helping us refine the questionnaire to improve clarity and accessibility.

The cognitive testing sample included individuals aged 50 and above, deliberately selected to reflect a range of demographic characteristics. Of the 15 participants, there was a near-even split in age groups, with five participants aged 50-59, four aged 60-69, and six aged 70 or older. In terms of gender representation, the group included five males, nine females, and one individual identifying as other. Rather than aiming for statistical representativeness, we intentionally selected a diverse sample to ensure that the questionnaire would be accessible and relevant to individuals with different characteristics, experiences, and backgrounds. This approach allowed us to identify potential comprehension challenges across subgroups of older adults, strengthening the overall validity of the questionnaire and its applicability.

A key objective of cognitive testing was to evaluate the clarity and appropriateness of language for an older audience. Based on participant feedback, we refined complex wording, simplified response categories, and ensured that all questions were easily comprehensible.

Ethical approval for the ELSA wave 12 cognitive testing was given by the National Centre for Social Research Ethics Committee (ref P18148). Recruitment started on 21^st^ of October 2024 until 1^st^ of November, and fieldwork ran from 5^th^ to 15^th^ of November 2024.

Participants gave their verbal consent to take part in the study before taking part, consent was documented in a spreadsheet, furthermore during the interview they were asked if they are happy to continue with the cognitive interview and whether also agree to be recorded. The consent was witnessed by the interviewer.

### The questionnaire and results of the cognitive test

The ClimateMind50+ questionnaire is depicted in Figure 1. The results of the cognitive interviews summarised in Table 1. The short introduction to the module sets the context for the subsequent questions by briefly explaining the concept of climate change as long-term changes in temperatures and weather patterns, primarily caused by human activities such as burning fossil fuels. This introduction provides a simple and accessible explanation to ensure all respondents, regardless of their prior knowledge, understand the scope of the topic being addressed. The cognitive testing revealed that the introduction was well understood, generally aligning with what people were thinking of when considering climate change.

**Table 1.** Cognitive interviews results.

| **Original Questionnaire Item** | **Cognitive Interview Probe(s)** | **Participant Response** | **Issues Identified** | **Correction** | **Revised Questionnaire Item** |
| --- | --- | --- | --- | --- | --- |
| The ClimateMind50+ questionnaire introduction explaining climate change. | Was the introduction clear and easy to understand? | Participants generally understood the introduction and felt it aligned with their perception of climate change. | No major issues identified. | No changes needed. | Introduction remains unchanged. |
| 1.How would you rate your knowledge of climate change? | How did you interpret the response options? | Participants found the question clear and relevant. | No issues identified. | No changes needed. | Question remains the same. |
| 2.How worried are you about the impact of climate change for you personally, your family, and future generations? | Did the response options capture your level of concern? | Participants found "Somewhat worried" and "Not very worried" too similar. | Ambiguity in response options. | "Not very worried" was removed, leaving three categories: "Very worried," "Somewhat worried," and "Not at all worried." | Revised question with updated response categories. |
| 3.How worried are you about extreme weather events where you live? | Did this question make you think about your local area or climate change in general? | Participants found "Somewhat worried" and "Not very worried" too similar. Some participants considered extreme events globally rather than locally. | Ambiguity in response options. Wording led to a broader interpretation than intended. | As for Question 2. The question was revised to explicitly refer to the local area. | Revised question with updated response categories. "Thinking about where you live right now, how worried are you about extreme weather events?" |
| 4.Which of the following statements apply to you regarding preparedness for extreme weather? | Were the response options clear? | The response "I can use nearby public spaces" was unclear to some respondents. | Lack of clarity in wording. | Reworded as "I can rely on the help of the local council" for better clarity. | Revised question with clearer response options. |
| 5.Have you made any changes to your lifestyle due to environmental concerns? | Did the response categories capture your actions accurately? | Participants found the frequency-based response options confusing. | Timeframe for actions not to be restricted. Overly complex response structure. | New response options: "Not at all" "Occasionally, when convenient" "Often, even if it requires effort" "Always, even if it requires significant sacrifice" | Timeframe for actions adjusted from "in the past two years" to "ever". Revised question with extensive response format and timeframe changed to "ever". |
| 6.Have you ever taken any of the following climate actions? | Were you able to accurately reflect your engagement in climate actions? | Participants found the timeframe restrictive. | The specified timeframe limited responses. Suggested to change the response options. | The timeframe was removed, and response options were streamlined. | Revised question with broader timeframe and simplified responses. Response options changed to Ye/No. |

Question 1 of the survey focuses on how participants would rate their knowledge of climate change and is designed to evaluate their self-perceived understanding of the topic. This question helps identify the level of confidence respondents have in their knowledge, which is essential for tailoring educational interventions and communication strategies to address any gaps or misconceptions. The cognitive testing revealed that this question was well understood, and no changes were needed.

Question 2 assesses participants’ levels of worry about the impact of climate change for them personally, their family and friends, and future generations. It aims to capture the recognition of its potential risks and consequences on themselves and their social circles. Question 3 then seeks to assess perceptions of risk and concern regarding specific climate-related events that may directly impact participants’ lives and local environments. The question asks participants how worried they are about the occurrence of various extreme weather events where they currently live, including extended droughts, severe flooding, extreme heat, storms with strong winds, hail or snow, wildfires (and related smoke or pollution), and prolonged or extremely cold winters. Questions 2 and 3 were assessed on a 4 response categories during the cognitive testing with responses as 1. Very worried, 2. Somewhat worried, 3. Not very worried and 4.

Not worried at all. There were a number of mentions that “Somewhat worried” and “Not very worried” were similar, we therefore removed the option “Not very worried” and kept the questions with 3 response categories instead. Furthermore, the wording of question 3 was changed from “How worried are you that any of the following extreme weather events might happen where you live right now?” to the current format as some respondents appeared to be thinking about these extreme events more generally as opposed to where they live now.

Question 4 asks participants to consider their preparedness for extreme weather events by selecting applicable statements regarding their reliance on support systems, the safety of their home, and their level of personal preparation. The options include relying on help from family, friends, neighbours, or the local council; confidence in the safety and comfort of their home; acknowledging a lack of preparation; or feeling that no preparation is necessary. This question helps to assess participants’ perceived readiness for extreme weather events and highlights any potential gaps in resources, awareness, or support networks that could impact their resilience. Following cognitive testing, the response option “I can use nearby public spaces” was rephrased to ”I can rely on the help of the local council”, as respondents found the former to be unclear. In countries where local councils do not exist, this response can be adapted to use contextually relevant wording to ensure clarity and applicability.

Question 5 is designed to assess respondents’ environmentally sustainable lifestyle practices and the changes they have made due to their concern for the environment. It consists of a list of specific actions that individuals may have taken to reduce their environmental impact, with a simple “Yes” or “No” response for each item. Respondents are also provided an open-ended section to describe other environmentally sustainable actions they may have taken, allowing for a broader understanding of individual contributions to environmental sustainability. This question helps capture both specific and diverse behaviours linked to environmental concern. This question required the most substantial revisions following cognitive testing to enhance its clarity and usability. Initially, the response categories measured the frequency of actions taken, but this was found to be overly complex and potentially confusing for respondents.

Therefore we further consulted with experts who suggested modifying the responses so that information on how often they had done each of the changes, and at what cost to them. The examples provided for each action were also significantly reduced, as the initial list was judged to be excessive and overwhelming, detracting from the question’s focus. Furthermore, the timeframe for considering these actions was adjusted from “in the past two years” to “ever,” as respondents indicated that a longer timeframe was more appropriate for capturing meaningful changes in behaviour, particularly among older populations. These changes collectively aimed to streamline the question and ensuring it effectively captured the intended data.

Question 6 aims to explore respondents’ engagement in various climate-related actions, reflecting their level of involvement and commitment to addressing environmental issues. Respondents are asked to select all applicable actions from a predefined list. Additionally, respondents can specify other climate-related activities they have undertaken in an open-ended section or indicate that they have not participated in any of the listed actions. During cognitive testing, this question was well understood by respondents; however, as in Question 5, the specified timeframe was dropped and replaced with “ever” to better capture the breadth of climate actions over a longer period of time. Furthermore, it was suggested that responses would be assessed only on a Yes/No scale rather than Always/Whenever possible/Never.

Lastly, it was recommended to add a couple of examples about activities undertaken to reduce waste.

Overall, cognitive testing provided key insights that enhanced the clarity and usability of the tool, such as refining response categories, simplifying complex wording, and adjusting timeframes to better reflect the experiences of older adults.

## Discussion

The main goal of this research was to introduce the ClimateMind50+ questionnaire, a newly developed tool for assessing key dimensions of climate-related knowledge, concerns, and adaptive behaviours among older adults, an area where measurement tools have been previously lacking.^2^ The aim of the questionnaire is to gather the experiences and viewpoints of this demographic, which when administered in surveys will help address a critical research gap and also provide evidence as to whether older adults are truly disengaged from climate issues and role they can play in promoting sustainable practices and contributing to climate change mitigation efforts.

Experts’ input led to refinements to ensure that the questionnaire effectively captures older adults knowledge, concerns, preparedness, and involvement in climate action while making it more accessible and meaningful for this demographic. We also reported detailed results of the cognitive testing process, which provided important insights into how older adults engage with the ClimateMind50+ questionnaire and highlighted areas where refinements improved clarity and accuracy. While participants generally understood the questions well, modifications were necessary to enhance accessibility, simplify response categories, and ensure relevance to this demographic. By prioritizing language appropriateness, we ensured that the questionnaire is accessible to a broad range of older adults, making it a practical tool for future research.

Significantly, the cognitive testing findings reinforced that while older adults could generally understand and engage with the questionnaire, certain refinements were necessary to ensure clarity and relevance. The removal of ambiguous response categories (e.g., simplifying the worry scale from four to three options) improved the accuracy of responses. Adjustments such as shifting behaviours-related questions to a lifetime (ever) timeframe rather than a recent-years window allowed for better recognition of long-term engagement. Additionally, the preparedness question was modified to clarify the role of personal readiness and external support systems, addressing initial confusion among respondents.

Future applications of this questionnaire can provide valuable data to inform policies and interventions that better support older populations in climate adaptation and mitigation efforts. By identifying key areas of concern, such as knowledge gaps, preparedness challenges, and engagement in climate action, the findings can support initiatives aimed at promoting adaptive strategies, improving access to resources, and integrating older adults into broader sustainability efforts.

## Strengths and Limitations

The ClimateMind50+ questionnaire presents several strengths. One key strength is its focus on an underrepresented population-older adults-who face unique vulnerabilities and challenges related to climate change but are often excluded from climate-related research. By specifically tailoring the questionnaire for individuals aged 50 and above, the questionnaire ensures that their perspectives, concerns, and adaptive capacities are adequately captured.

Additionally, the questionnaire comprehensively addresses multiple dimensions of climate change engagement, including knowledge, worries, preparedness, sustainable behaviours, and involvement in climate action. Our cognitive testing yielded valuable insights, showing that cognitive interviews are a useful tool for uncovering challenges such as clarity, understanding, ambiguity, memory load, timeframe considerations, missing response options, misleading instructions, and the overall relevance of questionnaire items.^24^

There are some limitations to this work. The sample size for cognitive testing was relatively small (n=15), which, while sufficient for identifying comprehension issues and refining the questionnaire, does not provide a statistically representative validation of the tool. Instead, the sample was intentionally selected to reflect diverse demographic and experiential backgrounds rather than broad representativeness. It is our hope that by administering the questionnaire in the main ELSA sample, as well as to larger and more diverse samples, will further validate the questionnaire and assess its applicability across different cultural and socioeconomic contexts. Additionally, the study relies on self-reported measures, which may be subject to recall or social desirability biases, particularly in areas such as climate-related behaviours and preparedness. However, the structured format of the questionnaire helps to minimize these biases by providing clear and standardized response options.

## Conclusion

The ClimateMind50+ questionnaire is an effective tool for measuring several aspects of climate change that are particularly relevant for individuals aged 50 and above. It enables the collection of critical data to understand the relationship between climate change and healthy aging. Designed to be quick to administer, the questionnaire offers flexibility in its delivery- whether face-to-face, by telephone, or through self-administration online or on paper-thanks to its simplicity and ease of completion. By addressing dimensions such as knowledge, worries, preparedness, behaviours, and engagement, the administration of the ClimateMind50+ will allow collection of climate change perspectives, empowering researchers and policymakers to craft informed strategies for mitigating climate risks while promoting sustainable and healthy aging practices.

This study contributes to the growing literature on aging and climate change by providing a structured tool specifically designed to capture climate-related concerns and actions in older adults. While previous research has often overlooked this demographic, the questionnaire helps bridge an existing gap by offering a standardized measure to assess climate knowledge, preparedness, and behaviours in this population.

Future research should focus on implementing the ClimateMind50+ questionnaire in large, more diverse populations to explore variations in climate engagement across different socioeconomic backgrounds and geographic regions. Additionally, longitudinal studies could examine how climate attitudes and behaviours evolve with age and changing environmental conditions.

## Data Availability

All data produced in the present work are contained in the manuscript

## Acknowledgement

We would like to thank the National Centre for Social Research, and in particular Lina Lloyd, for conducting the cognitive testing and providing extensive support in the development of the questionnaire. Their contributions have been invaluable to this project.

## References

1. Prina M, Khan N, Akhter Khan S, et al. Climate change and healthy ageing: An assessment of the impact of climate hazards on older people. J Glob Health. 2024;14:04101.

2. Yu-Tzu Wu, Matthew Prina, Paola Zaninotto, Climate change and healthy aging: What are the existing data in aging studies?, Innovation in Aging, 2025;, igaf008, 10.1093/geroni/igaf008

3. Ayalon L, Roy S. The Perceived Contribution of Older People to Climate Change Impact, Mitigation, and Adaptation: Measurement Development and Validation. Innov Aging. 2023 Sep 9;7(8):igad095. doi: 10.1093/geroni/igad095. PMID: 37841578; PMCID: PMC10576513.

4. 4. Kaiser, F. G. (2020). GEB-50. General Ecological Behavior Scale [Test description, questionnaire in German and English]. In Leibniz Institute for Psychology (ZPID) (Ed.), Open Test Archive. Trier: ZPID. 10.23668/psycharchives.4489

5. Steptoe A, Breeze E, Banks J, Nazroo J. Cohort profile: the English Longitudinal Study of Ageing. Int J Epidemiol. 2013;42(6):1640–8. doi:10.1093/ije/dys168. PMID:23143611; PMCID:PMC3900867.

6. Rowe JW, Kahn RL. Successful aging 2.0: Conceptual expansions for the 21st century. J Gerontol B Psychol Sci Soc Sci. 2015;70(4):593–6.

7. Pillemer KA, Nolte J, Cope MT. Promoting climate change activism among older people. Generations. 2022;46(2):1–16.

8. Hofer SM. Integrative data analysis and harmonization of longitudinal studies of aging and dementia. Innov Aging. 2017;1(suppl 1):1275.

9. Sanchez-Niubo A, Egea-Cortés L, Olaya B, et al. Cohort profile: The Ageing Trajectories of Health - Longitudinal Opportunities and Synergies (ATHLOS) project. Int J Epidemiol. 2019;48(4):1052–3i. doi:10.1093/ije/dyz077.

10. Lee J, Phillips D, Wilkens J, Gateway to Global Aging Data Team. Gateway to Global Aging Data: Resources for cross-national comparisons of family, social environment, and healthy aging. J Gerontol B Psychol Sci Soc Sci. 2021;76(suppl 1):S5–16.

11. Poortinga W. Review of environmental attitudes and behaviour questions in the Understanding Society survey. Understanding Society Working Paper 2022-03. Colchester (UK): University of Essex; 2022.

12. Higginbotham N, Connor L, Albrecht G, et al. Validation of an Environmental Distress Scale. EcoHealth. 2006;3:245–54.Office for National Statistics. Public attitudes to the environment and the impact of climate change, Great Britain: November 2021. GOV.UK. Published November 5, 2021. Accessed November 30, 2024. Available from: https://www.gov.uk/government/statistics/public-attitudes-to-the-environment-and-the-impact-of-climate-change-great-britain-november-2021

13. UK Department for Business, Energy & Industrial Strategy. Public attitudes tracking survey collection. Accessed November 30, 2024. Available from: https://www.gov.uk/government/collections/public-attitudes-tracking-survey

14. Irish Environmental Protection Agency. Climate change in the Irish mind. Accessed November 30, 2024. Available from: https://www.epa.ie/environment-and-you/climate-change/what-is-epa-doing/national-dialogue-on-climate-action/climate-change-in-the-irish-mind/

15. Kaiser FG. GEB-50: General Ecological Behavior Scale [verfahrensdokumentation, fragebogen deutsch und englisch]. In: Open Test Archive. Trier: Leibniz-Zentrum für Psychologische Information und Dokumentation (ZPID); 2020. doi:10.23668/psycharchives.3453.

16. Leßmann O, Masson T. Sustainable consumption in capability perspective: Operationalization and empirical illustration. J Behav Exp Econ. 2015;57:64–72.

17. Clayton S, Karazsia BT. Development and validation of a measure of climate change anxiety. J Environ Psychol. 2020;69:101434. doi:10.1016/j.jenvp.2020.101434.

18. Centre for Climate Change and Social Transformations. CAST website. Accessed November 30, 2024. Available from: https://cast.ac.uk/

19. Poortinga W, Demski C, Steentjes K. Generational differences in climate-related beliefs, risk perceptions and emotions in the UK. Commun Earth Environ. 2023;4:229.

20. UK Department for Energy Security and Net Zero. Climate change and net zero: Public awareness and perceptions. Published June 24, 2021. Accessed November 30, 2024. Available from: https://www.gov.uk/government/publications/climate-change-and-net-zero-public-awareness-and-perceptions

21. Arnold A, Böhm G, Corner A, et al. European perceptions of climate change: Socio-political profiles to inform a cross-national survey in France, Germany, Norway, and the UK. Oxford: Climate Outreach; 2016.

22. Steentjes K, Demski C, Seabrook A, et al. British public perceptions of climate risk, adaptation options and resilience (RESiL RISK): Topline findings of a GB survey conducted in October 2019. Cardiff: Cardiff University; 2020.

23. Balza, J. S., Cusatis, R., McDonnell, S. M., Basir, M. A., & Flynn, K. E. (2022). Effective questionnaire design: How to use cognitive interviews to refine questionnaire items. Journal of neonatal-perinatal medicine, 15(2), 345–349. 10.3233/NPM-210848

